# COVID-19: How to Relax Social Distancing If You Must

**DOI:** 10.1101/2020.04.29.20085134

**Authors:** Daniel Duque, David P. Morton, Bismark Singh, Zhanwei Du, Remy Pasco, Lauren Ancel Meyers

## Abstract

Following the April 16, 2020 release of the Opening Up America Again guidelines for relaxing COVID-19 social distancing policies, local leaders are concerned about future pandemic waves and lack robust strategies for tracking and suppressing transmission. Here, we present a framework for monitoring COVID-19 hospitalization data to project risks and trigger shelter-in-place orders to prevent overwhelming healthcare surges while minimizing the duration of costly lockdowns. Assuming the relaxation of social distancing increases the risk of infection ten-fold, the optimal strategy for Austin, Texas—the fastest-growing large city in the US—will trigger a total of 135 [90% prediction interval: 126–141] days of sheltering, allow schools to open in the fall, and result in an expected 2929 deaths [90% prediction interval: 2837–3026] by September 2021, which is 29% the annual mortality rate. In the months ahead, policy makers are likely to face difficult choices and the extent of public restraint and cocooning of vulnerable populations may save or cost thousands of lives.

## 1 Background

As of April 28, 2020, the COVID-19 pandemic continues to spread worldwide and has claimed at least 211 thousand lives (*1*). To avert unmanageable surges in COVID-19 hospitalizations, state and local policy makers across the US have implemented shelter-in-place orders to enforce social distancing. For example, the city of Austin, Texas, enacted the *Stay Home – Work Safe Order* (SHWSO) (*2*) on March 24, 2020, requiring individuals to stay at home except for certain essential activities. The decision to do so was partially based on model projections provided by the University of Texas at Austin. Three weeks later, a second order was issued, requiring cloth face coverings in public spaces. Under mounting pressures to relieve the economic and societal stresses of shelter-in-place orders, the US White House and Centers for Disease Control and Prevention (CDC) issued *Opening Up America Again* on April 16, 2020, which is a three-phased plan for relaxing such restrictions around the country (*3*). Texas followed suit, declaring that the state would begin returning to work, entertainment and commerce on May 1, 2020 (*4*).

In the absence of prophylactic and therapeutic countermeasures, non-pharmaceutical interventions are the only option for mitigating pandemic morbidity and mortality. Measures such as closures of schools and non-essential businesses, prohibitions on public gatherings, requiring social distancing, restricting travel, along with ordering face covering, frequent hand washing, surface cleaning, and staying at home when sick, can reduce both the frequency and risks of contacts between susceptible and infected people. During the early months of the 1918 influenza pandemic— the only comparably severe pandemic in recent history—aggressive social distancing proved critical to reducing mortality in the US (*5*). Despite the life-saving potential of social distancing measures (*6*, *7*), they are controversial (*8*) given their potentially large economic (*9*), societal, and mental health (*10*) costs. Two recent studies have projected pandemic resurgence if social distancing measures are relaxed prematurely (*11*, *12*), and others recommend the gradual relaxation of social distancing measures only when hospitals are no longer overburdened to balance expected public health risks and economic strain (*13–15*).

To this end, the goals of this article are threefold. First, we present a conceptual and quantitative framework that clarifies COVID-19 policy options for mitigating risk in the wake of the *first pandemic wave*. Second, we apply the framework to derive optimal triggers for initiating and relaxing shelter-in-place orders to minimize the number of days of costly social distancing while ensuring that COVID-19 hospitalizations do not exceed local capacity. Finally, we demonstrate the incontrovertible importance of sheltering vulnerable populations to reduce the burden of COVID-19. The impact of future social distancing policies will depend on public adherence, which is highly unpredictable. There are roughly two possible futures. In the first future, the pandemic is held at bay through a combination of public willingness to sustain extreme social distancing despite its costs and a ramping up of testing, contact tracing and isolation to rapidly contain emerging clusters. In the other future, a relaxation in social distancing or insufficient containment resources allow a second pandemic wave to emerge. For a policy maker facing the latter scenario, either intentionally or unintentionally, our framework provides guidance for enacting short-term lock-downs based on trends in local hospitalization data to avert unmanageable hospital surges while minimizing social and economic disruption.

Our new optimization model, detailed in the Supplementary Materials, is designed to guide the relaxation of social distancing, if a policy maker must. To demonstrate, we derive optimal surveillance triggers for enacting and lifting temporary shelter-in-place orders in the Austin-Round Rock Metropolitan Area of Texas (henceforth Austin) with a high-risk population of 547,474 and total population of 2,168,316. The analyses presented herein were requested by the Austin mayor and county judge and are informing ongoing risk assessments, policy planning, and public messaging. We assume that the COVID-19 epidemic began with a single case in Austin on February 15, 2020 and that the March 24th SHWSO will reduce transmission by 95% through May 1st, as estimated from local hospitalization data (*16*). We project pandemic hospitalizations under various intervention scenarios through September 2021. Schools are assumed to remain closed from March 14 until August 18, 2020. After that, they can be reopened or closed in tandem with shelter-in-place orders (*16*). After May 1, we assume that the city is either in a *relaxed* state in which the transmission rate is partially but not fully reduced by limited measures and efforts to test, trace and isolate cases, or a *lock-down* state in which renewed shelter-in-place orders reduce transmission by 90% relative to baseline. We find simple triggers for issuing sheltering orders and estimate the impact of *cocooning* vulnerable populations, that is, maintaining a 95% reduction in transmission to high-risk individuals. These findings provide actionable insights for other metropolitan areas where shelter-in-place orders have curbed the first wave of the COVID-19 pandemic. Moreover, the framework can incorporate any dynamic model of COVID-19 transmission to support similar planning throughout the US.

## 2 Results

All of our results are based on simulating variable levels of social distancing using a data-driven model for COVID-19 transmission and healthcare needs in the Austin, Texas metropolitan area (*16*). Austin can toggle between a *relaxed* state in which transmission is reduced by 40% relative to the baseline transmission rate estimated prior to schools closing on March 14 and a *lock-down* state in which transmission is reduced by 90%. The relaxed state does not fully rebound to baseline transmission under the assumption that testing-based containment and voluntary social distancing will partially mitigate risks. Further analyses for other degrees of relaxation, ranging from a 20% to 80% reduction in transmission, are provided in the Supplementary Materials. All projections end in September 2021, which is an optimistic time horizon for the development and distribution of prophylactic or therapeutic medical countermeasures (*17*).

To evaluate and optimize intervention policies, we compare two outcome measures. First, we measure the total number of days of lock-down (i.e., shelter-in-place) until Sep 30, 2021 as a proxy for the economic and societal costs of the policy, depicted by gray shading in Fig. 1. Second, we determine the probability of exceeding hospital capacity as a proxy for the public health risks of the policy, indicated when the red hospitalization curves surpass the red capacity line in Fig. 1.

**Fig. 1.**
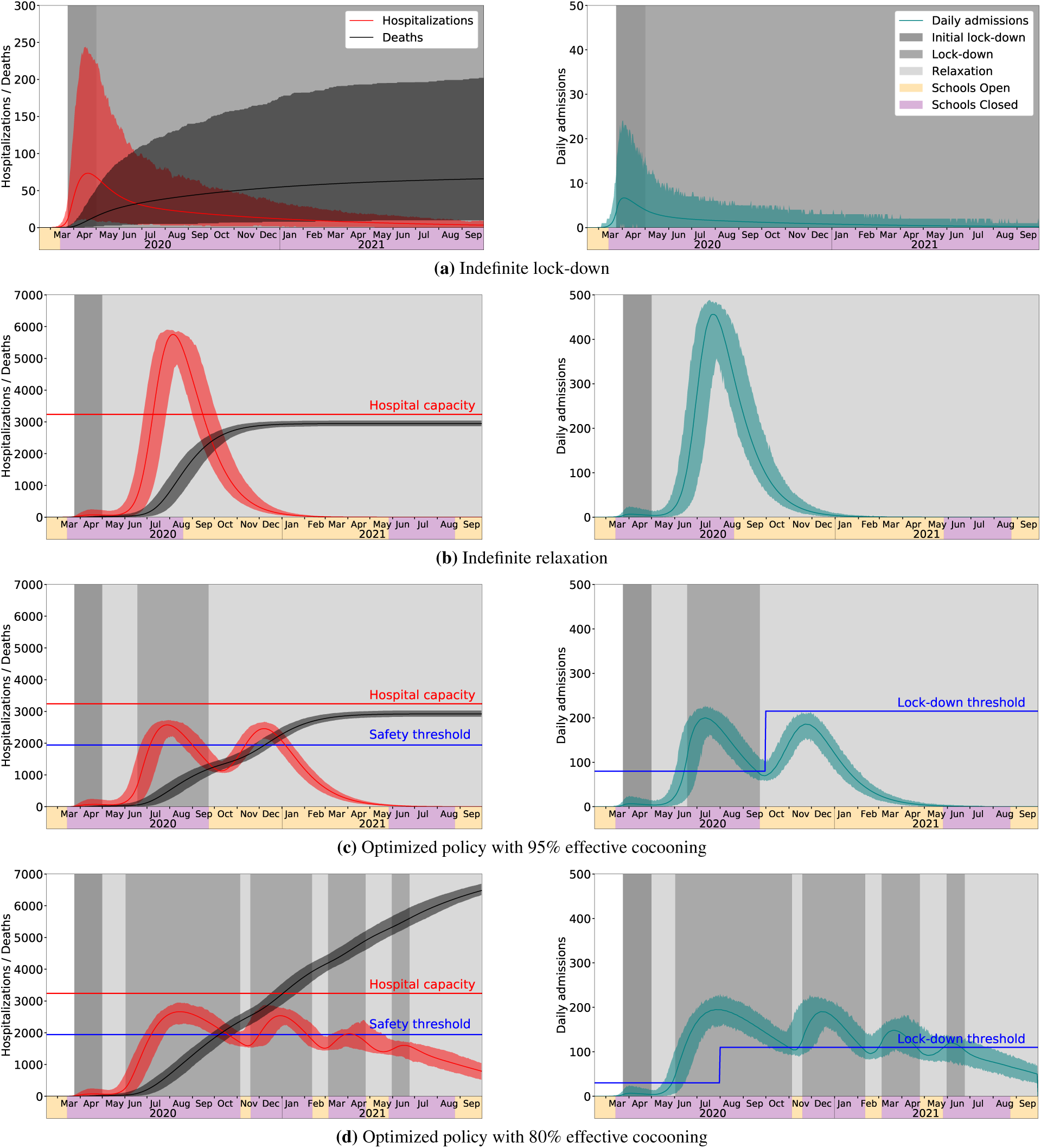
Projections for COVID-19 hospitalizations and deaths in Austin, Texas metropolitan area under baseline and optimized policies for initiating and relaxing social distancing measures. Left-hand panels show daily hospitalizations and cumulative deaths. COVID-19 surge capacity in Austin is approximately 3240 beds (red Une). Optimized strategies can relax lock-downs when total hospitalizations drop below a safety threshold of 60% capacity (blue line). Right-hand panels show daily hospital admissions. Optimized strategies use a stepped threshold: lock-downs are enacted when daily admissions (seven-day rolling average) surpass the threshold and are relaxed when admissions decline below the threshold, if hospitalizations are below 60% capacity. (a) The lock-down continues through September 2021, resulting in a 90% reduction in transmission, along with vigilant cocooning of vulnerable populations (95% effective), and school closures. (b) The lock-down is relaxed on May 1, 2020. Thereafter, transmission is reduced by 40%, schools open in mid-August 2020, and 95% effective cocooning of vulnerable populations is maintained through September 2021. Hospitalizations are expected to grossly overrun capacity. (c) Adaptive lock-downs triggered when hospital admissions cross optimized thresholds, assuming 95% effective cocooning of vulnerable populations. The thresholds minimize the expected days of lock-down while ensuring hospital capacity is not exceeded with high probability. (d) Adaptive lock-downs when effectiveness of cocooning drops to 80%. Even under an optimized solution, expected deaths and days in lock-down both more than double, relative to cocooning at 95%. In all graphs, solid curves correspond to the point forecast and shaded regions give 90% prediction intervals based on 300 stochastic simulations.

We first project COVID-19 hospitalizations in the extreme scenario that the city maintains a 90% reduction in local transmission indefinitely through a combination of aggressive social distancing, transmission-reducing precautions and proactive testing, contact tracing, and isolation (Fig. 1a). The analysis assumes that schools remain closed, and cocooning of high-risk populations reduces their risk of infection by 95% rather than 90%. Under this policy, we would not expect a second wave to emerge during the model horizon (Fig. 1a). Cumulative deaths would be expected to slowly climb to 81 [90% prediction interval: 10–202]. This scenario costs a year and a half (555 days) of lock-down.

In the other extreme, consider the scenario in which Austin permanently relaxes social distancing on May 1st, while continuing to cocoon high-risk populations and opening schools on August 18th (Fig. 1b). While this policy requires a lock-down for only the initial 38-day period prior to May 1st, we would expect a catastrophic surge in hospitalizations that exceeds the local capacity by 80% during July–September 2020, resulting in an expected 23,075 [90% prediction interval: 22,409–23,741] patients not receiving critical care. Without accounting for the excess mortality during this period, which could be considerable, we would expect at least 30-fold higher COVID-19 mortality relative to the indefinite lock-down scenario, with deaths reaching 2957 [90% prediction interval: 2868–3040] by September 2021. Under this policy, we expect two epidemic waves during the model horizon, with the second large wave peaking in the late summer of 2020.

Assuming that the first scenario is unattainable and the second scenario unacceptable, we seek alternative policies that limit the number of days in lock-down while preventing COVID-19 health-care surges beyond local capacity. Based on our decision-support efforts for the city of Austin and potential biases in confirmed case count data across the US, we conjecture that local hospitalization data will be a more reliable indicator of transmission intensity and future hospital surges. Our best policies track daily COVID-19 hospital admissions and daily total hospitalizations across the city and trigger the initiation and relaxation of lock-down periods when admissions cross predetermined thresholds.

Specifically, we formulate and solve a stochastic optimization problem that selects daily hospitalization triggers and recommends reinstatement and relaxation of lock-down periods as follows:

1. reinstate the lock-down—corresponding to a 90% reduction in transmission—when the seven-day average of daily hospital admissions exceeds the trigger; and
2. release the lock-down—corresponding to a 40% reduction in transmission—when both: (a) the seven-day average of daily hospital admissions drops below the trigger, *and* (b) city-wide hospitalizations (heads in beds) are below a fixed factor (60%) of surge capacity for COVID-19.

If randomized testing becomes available at sufficient scale, we could similarly determine triggers based on testing rather than hospitalization data, and thereby gain earlier indications of a rising or declining threat.

To keep estimated hospitalizations within capacity, we use the square-root staffing rule from queueing theory (*18*). This rule maintains a high probability (we use ≥ 0.9999) that a single arrival in steady state does not have to wait for service, and yet servers are highly utilized. Our “servers” are hospital beds, along with necessary healthcare providers and equipment. We assume that 80% of Austin’s hospital beds are available for COVID-19 patients. We require that the square-root staffing rule hold, under a point forecast for daily COVID-19 hospitalizations. In addition we simulate, and optimize with respect to, 300 sample paths of the epidemic, taking into account both macro- and micro-level stochastics, with details in the Supplementary Materials. We ensure that the probability of exceeding hospital capacity within the time horizon is at most 0.01. With these constraints in place, we select triggers to *minimize* the expected number of days of lock-down. Minimizing lock-down acknowledges social and economic pressures to relax stringent measures.

Given the hospital capacity in the Austin, Texas metropolitan area we recommended a simple, yet robust, strategy with two fixed thresholds: 80 daily COVID-19 hospital admissions prior to September 30, 2020, and 215 thereafter, as indicated by the blue step function in the right panel of Fig. 1c. We optimized these two values as well as the date of the transition. Under the point forecast for the pandemic, 135 days of lock-down are required and hospitalizations remain safely below capacity. Stochastic simulation yields a mean of 135 days [90% prediction interval: 126–141]. The projected mortality is substantial, with a mean of 2929 deaths [90% prediction interval: 2837–3026], which is again over 30 times larger than the baseline scenario of indefinite lock-down (Fig. 1a). While the other baseline scenario of indefinite relaxation projected similar COVID-19 mortality (Fig. 1b), it produces a catastrophic surge in hospitalizations, and those projections do *not* account for excess mortality caused by inadequate healthcare resources during the July-September 2020 surge period.

These projections assume an ambitious policy of cocooning vulnerable populations with a 95% level of effectiveness. If cocooning only attains an 80% reduction in transmission risk, then we would expect far greater numbers of hospitalizations and deaths. Under this scenario, the optimal policy requires lower thresholds for enacting lock-downs: 30 daily COVID-19 hospital admissions prior to July 31, 2020 and 110 thereafter (Fig. 1d, Table 1). *Leaky* cocooning can substantially undermine containment. In this case, the optimal strategy for managing hospital surge requires multiple periods of lock-down totaling about 350 days and more than doubling expected mortality.

**Table 1:** Projected days of lock-down, probabilities of exceeding hospital capacity and COVID-19 mortality under the optimized strategies with 95% and 80% effective cocooning of vulnerable populations. The third and fourth columns correspond to Fig. 1c and Fig. 1d, respectively. Note that Fig. 1 shows 90% prediction intervals (i.e., 5%-95%) for hospitalizations based on 300 simulations, and the last two rows of the table correspond to more extreme events.

| Scenario |  | Cocooning at 95% | Cocooning at 80% |
| --- | --- | --- | --- |
| Days of lock-down | Mean | 135 | 346 |
|  | Median | 135 | 347 |
|  | 5%-95% PI | (126-141) | (333-360) |
| Cumulative deaths by Sep 30, 2021 | Mean | 2929 | 6527 |
|  | Median | 2646 | 6532 |
|  | 5%-95% PI | (2837-3026) | (6338-6690) |
| Risk Group | Age Group | % deaths |  |
| Low risk | 0-4y | 0.03 | 0.02 |
|  | 5-17y | 0.20 | 0.08 |
|  | 18-49y | 9.01 | 3.60 |
|  | 50-64y | 18.23 | 6.97 |
|  | 65y+ | 4.43 | 6.12 |
| High risk | 0-4y | 0.00 | 0.02 |
|  | 5-17y | 0.10 | 0.09 |
|  | 18-49y | 6.04 | 5.82 |
|  | 50-64y | 26.86 | 28.32 |
|  | 65y+ | 35.06 | 48.95 |
| Probability of exceeding % of hospital capacity | 60% | 1.000 | 1.000 |
|  | 70% | 0.997 | 1.000 |
|  | 80% | 0.803 | 0.973 |
|  | 90% | 0.020 | 0.197 |
|  | 100% | 0.000 | 0.007 |

We conducted sensitivity analyses to assess the robustness and limitations of the optimized triggers. The proposed triggers are relatively robust to weaker social distancing during relaxation periods, e.g., if transmission is only reduced by 20% rather than 40%. However, the proposed triggers are *not* robust to leaky cocooning. We analyze the relative merits of policies with a constant lock-down threshold to the horizon, relative to having two distinct thresholds, as presented here. We show the importance of optimizing trigger thresholds: conservative triggers significantly increase the duration of lock-down periods, while loose triggers result in hospital capacity being overrun. See Supplementary Materials for details.

## 3 Discussion

A significant relaxation of social distancing in the absence of a comprehensive program for testing, contact tracing and isolation will likely lead to future waves of the COVID-19 pandemic in US cities. Even if policy-makers extend lock-down periods, lack of public willingness to comply might undermine their efficacy. Thus, planning for future relaxations is paramount to averting unmanageable surges in COVID-19 hospitalizations. Carefully designed strategies for triggering future shelter-in-place measures can mitigate the impact on the community’s healthcare system while minimizing economic and societal costs.

Our framework clarifies a key decision facing city and state leaders in the wake of the first wave of COVID-19 transmission—when to enact and relax social distancing measures should the epidemic rebound. We posit a simple strategy for measuring and responding to future surges in hospitalizations—enact and then relax temporary lock-downs when daily hospital admissions climb above and eventually recede below a predetermined, optimized threshold.

The optimal strategies derived for Austin, Texas provide three critical insights. First, data-driven optimization yields policies that are expected to protect against catastrophic hospital surges while requiring far fewer days of costly shelter-in-place measures than most sensible expert-designed strategies. For example, triggering lock-downs based on an arbitrarily chosen trigger of 50 new admissions per day should prevent hospitalizations from reaching capacity, but they are expected to require more than 150 additional days of lock-down, relative to the optimized trigger policy. However, implementing this trigger-based optimization framework requires continual review of daily hospital admissions and overall hospital utilization, as well as constant validation of transmission rates during lock-down and relaxation phases.

Second, under the plausible scenario that transmission rebounds to 60% of baseline (i.e., a 40% reduction), the best strategy for limiting lock-downs without undermining the healthcare system would likely trigger only one future lock-down in mid June following a steep increase in hospitalizations that surpasses the trigger of 80 new admissions per day (Fig. 1c). Hospitalizations would then be expected to peak and subside in late July, allowing relaxation of the lock-down by late September. The simultaneous release of the lock-down and start of a delayed 2020–2021 school year would fuel a third wave, which would be expected to be self-limiting, that is, subside without requiring a third lock-down period. This decline is driven by herd immunity, with an expected 79% of the population already infected and recovered by October 2021.

We emphasize that, while this strategy offers a practical balance between economic and health-care constraints, it is not designed to minimize morbidity and mortality and results in nearly 3000 expected deaths by September 2021. If we assume a similar COVID-19 mortality rate for the entire US, this extrapolates to over 450 thousand deaths, an order of magnitude higher than the annual mortality from seasonal influenza. We note, with concern, that this alarming projection assumes that high-risk populations maintain 95% effective social distancing through September 2021.

Finally, failing to vigilantly cocoon our vulnerable populations will significantly increase both the death toll and the requisite number of days in lock-down, even under the most efficient policy for keeping hospitalizations in check. Nursing homes and populations experiencing homelessness have both large proportions of high-risk individuals and living conditions that exacerbate the risks of COVID-19 transmission. Proactive measures to prevent COVID-19 introductions into these communities and to rapidly contain initial clusters is essential to effective cocooning but will require considerable forethought and resources, including additional trained staff and isolation facilities. In addition, providing incentives and support for high-risk members of the workforce to shelter-at-home will be critical.

While we believe that our qualitative findings are robust and provide actionable insights for navigating the challenges ahead, our quantitative findings are specific to Austin and are based on several simplifying assumptions. For example, we do not consider the impact of the 2020–2021 influenza season on surge capacity for COVID-19 cases. During the 2019–2020 influenza season, several Austin area hospitals neared their capacity. In reality, we cannot predict when or how much transmission will rebound from policy loosening or public fatigue. Yet, our optimal policies assume a specific and constant degree of relaxation. Our sensitivity analysis suggests that the derived policies are relatively robust to uncertainty regarding future transmission but not to a relaxation of cocooning.

Our simple threshold policies allow optimization using a relatively small discrete grid. This approach can be directly applied to other epidemiological simulation models, provided that they can incorporate both triggers and variable levels of social distancing, e.g., by adjusting transmission rates over time. Thus, modelers can broadly apply this framework to provide decision-support for COVID-19 responses in cities worldwide. In addition to tracking hospitalizations for triggering shelter-in-place orders, modelers will need to regularly estimate local transmission rates as policies and individual behavior evolve. Although daily COVID-19 hospitalization counts allow robust estimation of reductions in transmission (*16*), the signal is delayed by the roughly 10-day lag between infection and hospitalization. Integrating cell phone mobility data reflecting social distancing, as we do in our forecasting model (*19*, *20*), may improve the accuracy and timeliness of our estimates. Finally, September 2021 is a long horizon. In the months ahead, the likelihood and timeline for promising antiviral drugs and vaccines may become clearer. If such life-saving measures appear within reach, communities may have a renewed willingness to shelter-in-place that can be directly incorporated to designing new triggers for aggressive mitigation.

## Data Availability

All data is available in the main text or the supplementary materials.

## Acknowledgments

**Funding:** This work was supported by the National Institutes of Health under Grant NIH R01 AI151176 and by the U.S. Department of Homeland Security under Grant 2017-ST-061-QA0001. The views and conclusions contained in this document are those of the authors and should not be interpreted as necessarily representing the official policies, either expressed or implied, of the U.S. Department of Homeland Security.

## Data and materials availability

All data is available in the main text or the supplementary materials.

## Supplementary Materials for COVID-19: How to Relax Social Distancing If You Must

**This PDF file includes:**

Materials and Methods

Supplementary Analysis

Model Parameters

Figs. S1 to S5

Tables S1 to S14

### Material and Methods

We optimize the timing of when to initiate and relax local lock-downs (i.e., extreme social distancing measures) via triggers that monitor both daily new hospital admissions and total hospitalizations. A lock-down is triggered when a seven-day moving average of daily hospital admissions grows to exceed a *lock-down threshold*, which we specify for each day. Relaxation of a lock-down is triggered when: (a) the same moving average drops below the lock-down threshold and (b) the total hospitalizations are under a *safety threshold*. Here, lock-down corresponds to high social distancing (SD) —90% reduction in transmission in our instances—and a relaxation of SD means a lower level of SD—we use 40% reduction as a nominal value.

In addition to epidemiological constraints governing the transmission and severity of the virus, our optimization model selects triggers that ensure the aggregate daily arrival rate of new patients to hospitals is such that, with high probability, the demand for hospital beds does not exceed supply. We use the square-root staffing rule of (*18*) for an *M*/*M*/*s* queue with *s* servers. That rule says that under a service rate of *μ*, if the arrival rate, *λ*, is large then we require s 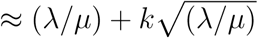 servers to maintain a high probability that the servers are highly utilized and yet an arrival does not have to wait for service. The rule is based on an approximation, which holds in steady state under independent and exponentially distributed interarrival times and service times, and the approximation is increasingly accurate as λ grows large. The value of parameter *k* determines the model’s probability that an arriving patient does not have to wait for a hospital bed, and we use *k* = 4, which corresponds to a probability of over 0.99997 for a single patient. We first describe a deterministic optimization model: Among all feasible triggers, the optimization model seeks triggers to minimize the number of days in lock-down.

To find the lock-down and safety thresholds, we formulate an optimization model that determines daily values for both thresholds, while hospitalizations and daily hospitalizations are characterized via an enhanced SEIR-style metapopulation model of disease transmission (*16*). The underlying epidemiological model comprises compartments for susceptible, exposed, infectious-asymptomatic, infectious-symptomatic, infectious-hospitalized, recovered, and deceased, which we denote by *S*, *E*, *IA*, *IY*, *IH*, *R*, and *D*, respectively. The population is partitioned into 10 groups comprised of all combinations of five age groups and two risk groups. Each group is represented with its own set of compartments (susceptible, exposed, etc.) so that in total the epidemiological model has 70 compartments. To formalize our model, let *X_t_* be a binary indicator variable that takes the value of one if a lock-down is in place on day *t*. The goal is to find thresholds *l_t_* and *r_t_* for all 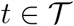 so that the sum of indicator variables is minimized while respecting hospital capacity and epidemiological dynamics.

#### Notation

Indices and Sets

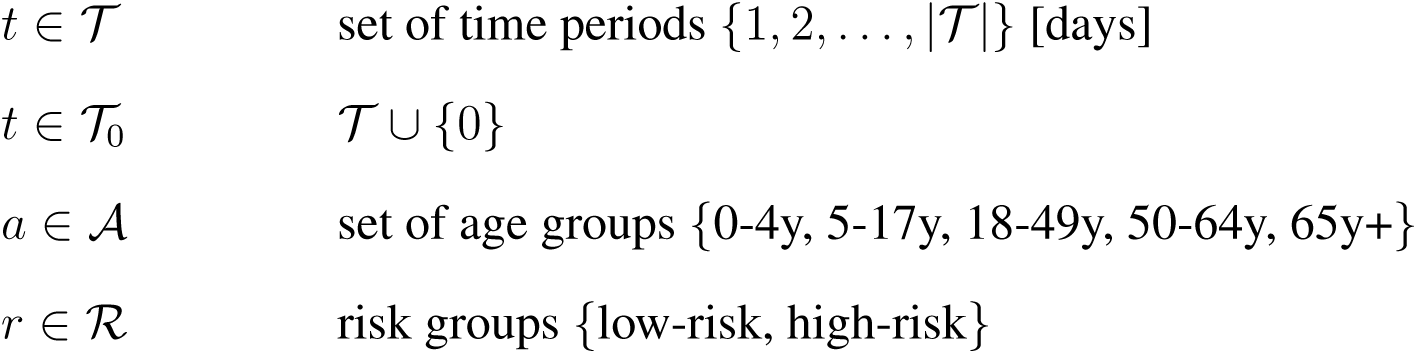

*Parameters*

*Epidemiological parameters:*

|  |  |
| --- | --- |
| $\beta$ | transmission rate |
| $\sigma$ | rate at which exposed individuals become infectious |
| $\tau$ | proportion of exposed individuals who become symptomatic |
| $\gamma_A$ | recovery rate from asymptomatic compartment |
| $\gamma_Y$ | recovery rate from symptomatic compartment |
| $\gamma_H$ | recovery rate from hospitalized compartment |
| $\eta$ | hospitalization rate after symptom onset |
| $YHR^{a,r}$ | percent of symptomatic infectious that go to the hospital for $a, r$ |
| $\pi^{a,r}$ | $\gamma_Y \cdot YHR^{a,r} / [\gamma_Y \cdot YHR^{a,r} + \eta(1 - YHR^{a,r})]$: rate adjusted proportion for $a, r$ |
| $\mu$ | rate from hospitalization to death; $\mu = \gamma_H$ in our instances |
| $HFR^a$ | percent of hospitalized that die for $a, r$ |
| $\nu^a$ | $\gamma_H \cdot HFR^a / [\gamma_H \cdot HFR^a + \mu(1 - HFR^a)]$: rate adjusted proportion for $a, r$ |
| $\phi_{a',r'}^{a,r,t,high/low}$ | expected number of daily contacts from $(a', r')$ to $(a, r)$ at $t$ under high or low SD |
| $N_{a,r}$ | population of age-risk group $a, r$ |
| $\omega_{A/E}^{a,r}$ | infectiousness of individuals in $IA$ and $E$ relative to $IY$ for $a, r$ |

Other parameters:

|  |  |
| --- | --- |
| $B$ | number of hospital beds for COVID-19 patients |
| $k$ | safety factor for square-root staffing rule |
| $\bar{\lambda}$ | largest $\lambda$ satisfying $\lambda/\mu + k\sqrt{\lambda/\mu} \leq B$ |

Variables

Epidemiological variables:

|  |  |
| --- | --- |
| $S_t^{a,r}$ | number of susceptible people of age group $a$, risk group $r$ at time $t$ [persons] |
| $dS_t^{a,r}$ | $S_t^{a,r} - S_{t+1}^{a,r}$ [persons] |
| $E_t^{a,r}$ | number of exposed people of age group $a$, risk group $r$ at time $t$ [persons] |
| $IA_t^{a,r}$ | number of infectious-asymptomatic people for $a, r, t$ [persons] |
| $IY_t^{a,r}$ | number of infectious-symptomatic people for $a, r, t$ [persons] |
| $IH_t^{a,r}$ | number of infectious-hospitalized people for $a, r, t$ [persons] |
| $R_t^{a,r}$ | number of recovered people for $a, r, t$ [persons] |
| $D_t^{a,r}$ | number of deceased people for $a, r, t$ [persons] |
| $IYIH_t$ | daily hospitalizations at time $t$ [persons/day] |
| $\overline{IYIH}_t$ | seven-day moving average of $IYIH_t$ [persons/day] |

Intervention variables:

|  |  |
| --- | --- |
| $X_t$ | 1 if lock-down (e.g., 90% SD) is applied at time $t$; 0 otherwise (e.g., 40% SD) |
| $l_t$ | the lock-down trigger for the daily hospitalization rate at $t$ |
| $r_t$ | the safety trigger for total hospitalizations |
| $\phi_{a',r'}^{a,r,t}(X_t)$ | expected number of daily contacts at $t$ determined by $X_t$ |

Epidemiological dynamics:

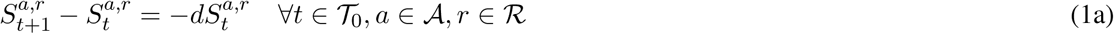

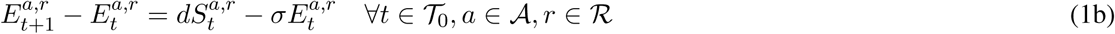

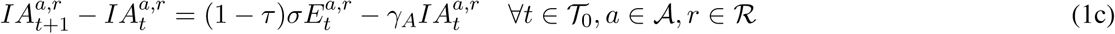

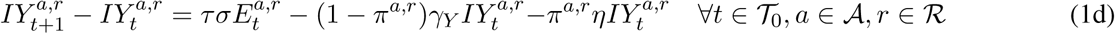

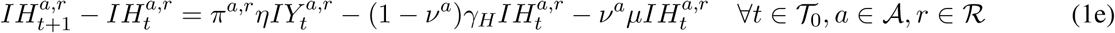

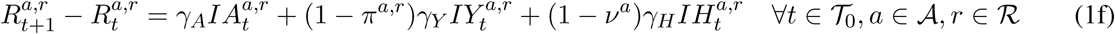

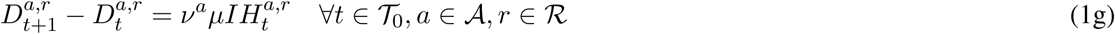

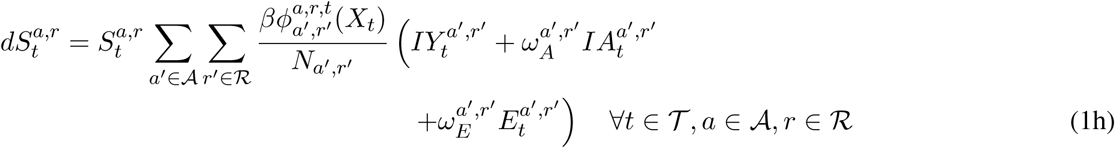

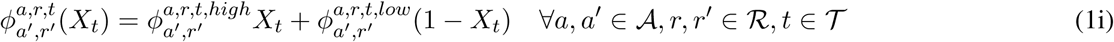

#### Initial conditions

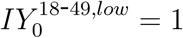, 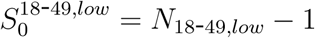, and 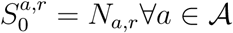, 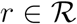.

The initial conditions specify a single infectious individual in the 18–49 age group with low risk. The age-risk groups are initialized with the rest of the population in their respective susceptible compartments. Equations (1a)-(1h) then provide a point forecast for the evolution of the disease. Variable *X_t_* ∊ {0,1} is a toggle that either selects the expected number of daily contacts according to the lock-down, or the relaxed, contact matrix: Parameter 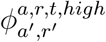 encodes the expected number of daily contacts on day *t* when we are in lock-down, i.e., high SD (e.g., 90% SD), and 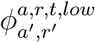 does the same for relaxed SD (e.g., 40% SD). These parameters are indexed by *t* because they capture whether school is currently open and if so, the school calendar; and, they further capture weekdays versus weekends and the level of cocooning, which can vary with time.

#### Threshold policy

The *X_t_* toggle is determined by the thresholds *l_t_* and *r_t_* using the following logic:

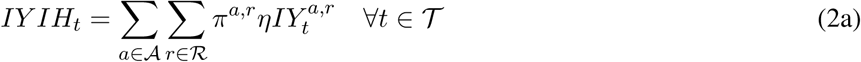

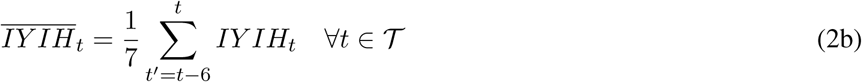

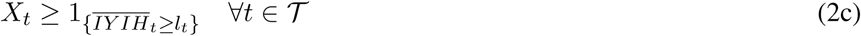

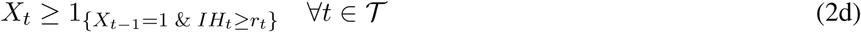

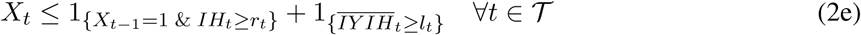

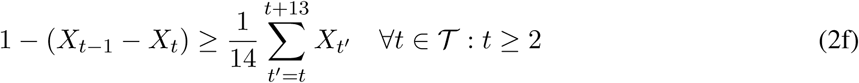

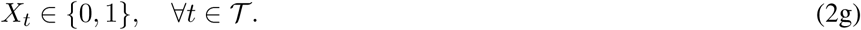

Systems (1) and (2) are coupled. Given *X_t_* we can specify a solution to system (1), and that solution defines the arrival rate to the community’s hospitals via (2a). The seven-day moving average of daily arrivals is defined in equation (2b). Given thresholds, constraint (2c) enforces a lock-down when that average is at or above the lock-down threshold, *l_t_*. If the average is below *l_t_* then constraint (2d) ensures we are still in lock-down if we were yesterday *and* the total number currently hospitalized exceeds the safety threshold. If neither of those events holds then inequality (2e) requires that we relax SD. To avoid impractical policies that lock-down and relax for short periods of time, inequality (2f) requires that a relaxation last at least fourteen days if enacted. Given the corresponding *X_t_*, equation (1i) again selects the corresponding daily-contact matrix, coupling the systems.

#### Deterministic optimization model

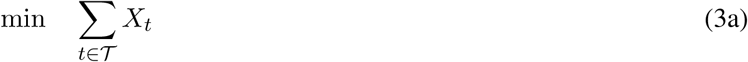

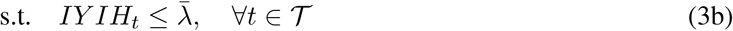

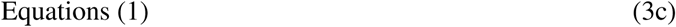

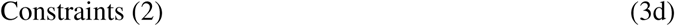

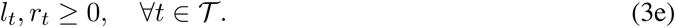

A solution to model (3) provides daily thresholds to guide both lock-down and relaxation of strict SD. Constraint (3b) enforces the square-root staffing rule, and the model’s objective is to minimize the number of days in lock-down.

Let (*l*, *r*) denote the vector of thresholds. Then we let:

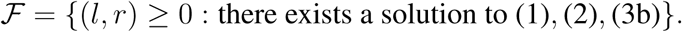

In other words, 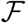 specifies the set of thresholds that ensure the arrival rate to hospitals is within the rate specified by the square-root staffing rule.

The deterministic finite-difference equations (1), or their differential equation counterparts, are approximations in which binomial random variables replace terms like 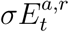; here the binomial random variable has parameter 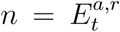 and *σ* serves as the “success” probability. This construct is pervasive throughout right-hand side terms in equations (1). In addition to these “micro” stochastics there are “macro” stochastics including modeling *σ* and γ*_Y_* and coupled parameters as random variables; see Table S7 for further details. As a result, except for (*l, r*) we can parameterize all variables by (*S*(*ω*)*,E*(*ω*)*, IA*(*ω*)*, IY(ω),IH*(*ω*)*, R*(*ω*)*, D*(*ω*)), where *ω* ∊ Ω indexes a sample path of the stochastic simulation. In this setting we solve the following model.

#### Stochastic optimization model

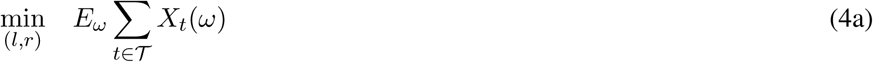

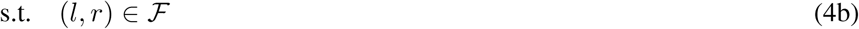

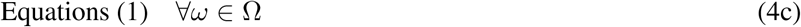

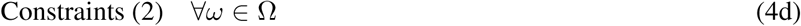

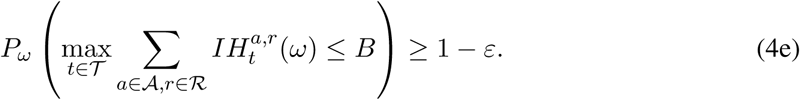

Constraint (4b) ensures the thresholds satisfy the square-root staffing rule for the best-guess (deterministic) epidemiological parameters. Constraints (4c)-(4d) track stochastic disease dynamics for each sample path. Constraint (4e) is a probabilistic constraint, which ensures that over the time horizon the maximum number of heads in beds is within hospital capacity for COVID-19 patients with high probability; we use *ε* = 0.01 in our computation. Subject to these constraints we minimize the expected number of days in lock-down in (4a). We do not compute exact expectations, but rather solve a sample average approximation of model (4) using Monte Carlo simulation to generate sample paths.

Having a different threshold each day is unrealistic for practical policy implementation. We seek a simpler policy, and so we restrict attention to policies with lock-down thresholds, *l_t_*, that can change only *once* during the planning horizon. This class of thresholds can be characterized with three parameters: (i) the threshold value up until the change, (ii) the threshold value after the change, and (iii) the timing of the change. In addition, in our implementation we required a single safety threshold *r* = *r_t_* for the entire horizon.

The deterministic model (3) can be formulated as a nonlinear mixed-integer program, and model (4) is a stochastic nonlinear mixed-integer program. In its general form, even the deterministic model is challenging to solve with commercial solvers, in part, because of the bilinear terms involving products of continuous variables that appear in constraint (1h). (These are further multiplied by binary variables, but those can be linearized exactly.) However, the class of policies just specified simplifies the task of finding an optimal solution, which involves just four decision variables: three associated with *l_t_* and one associated with *r_t_*. We optimize over the space of these variables as follows: For the lock-down threshold we parameterize the search on a grid, where the first two decision variables can take values between 0 and 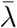 in steps of 10 patients per day, and the third decision variable can take values corresponding to the first day of every month. We similarly form a grid for the safety threshold *r*. To select an optimal policy, we first enumerate all possible tuples of the four decision variables and preclude those that do not satisfy the staffing rule defined in (3b), applying systems (1)-(2) so that contact matrices are selected on a daily basis according to how the triggers unfold in a deterministic simulation under best-guess parameter values. To obtain an optimal solution to model (4), we run stochastic simulations for each configuration, and then preclude those that do not satisfy inequality (4e), finally selecting the solution with the smallest number of expected days in lock-down.

The trigger-based policy is closed-loop, i.e., adaptive, in the sense that the timing of lock-down and relaxation depends on the sample path of the simulations. All results presented in the main text use 300 stochastic simulations to compute the threshold policy, and an independent set of 300 out-of-sample simulations to report performance of that policy.

We further investigated other classes of thresholds for the lock-down trigger, such as constant, linear, and quadratic thresholds as a function of time. While the step-function threshold for *l_t_* involves three parameters, a constant threshold is parameterized with one value, a linear threshold with two values (intercept and slope), and a quadratic threshold with three parameters (where the third parameter is the coefficient of the quadratic term). Using such restricted classes of thresholds provides policies that are both simple to implement and simple to optimize via simulation for all configurations on a discretized grid. This observation is important because it allows modelers to use our optimization framework with their own simulation model, provided that they can incorporate the dynamics of the triggers in their model (e.g., adjusting the contact matrix matrix over time).

Tables S7 and S8 give numerical values and probability distributions for the epidemiological parameters and hospitalization parameters that we use. Table S6 shows how overall age-risk group contact matrices are computed, as a function of time, based on individual contact matrices, which involve *home*, *school*, *work*, and *other* and further based on school closures, cocooning, and social distancing decisions. The specific contact matrices are then given in Tables S9-S12. After reporting additional analysis of trigger-based policies, the next section also details estimates of age-risk populations.

### Supplementary Analysis

We report additional results using the same format as the primary figure in the main text. For easy reference, Figure S1a repeats the results for the optimized two-level trigger for the baseline analysis; i.e., we assume 95% effective cocooning, and we toggle between 90% SD (lock-down) and 40% SD (relaxation). The daily hospitalization thresholds, and the timing of the transition, are optimized using model (4). We fix the safety trigger that prevents premature relaxation at 60% of hospital capacity. Part (b) of Figure S1 repeats this optimization, but instead restricts solutions to a *constant* rather than two-level threshold. Parts (c) and (d) show the results of *stress tests*, in which we assess the performance of the optimal strategies shown in parts (a) and (b) when relaxed social distancing reduces transmission by only 20% rather than 40%. Under the two-level trigger, the point forecast remains under hospital capacity but the distributional forecasts suggest a significant probability that hospitals will be at, or just beyond, capacity at the peak in July. Under the constant trigger, even the point forecast “mildly” exceeds hospital capacity.

Figure S2 performs an analogous stress test with respect to cocooning. First, we optimize for our baseline 90%-40% SD toggle, under the assumption that cocooning is 95% effective (as in Figure S1a). Then, we evaluate the performance under the assumption that cocooning is only 80% (rather than 95%) effective. Whereas our optimal policy is robust to variation in the overall efficacy of social distancing (Figure S1), it is highly sensitive to the efficacy of cocooning. This slight reduction in cocooning is expected to lead to catastrophic surges in hospitalizations, with capacity grossly exceeded in the summer and early fall of 2020. This reinforces one of our primary conclusions, that vigilant cocooning of vulnerable populations will be critical to preventing overwhelming healthcare surges and saving lives.

To quantify the benefit of deriving optimal triggers versus sensible expert-designed strategies, we consider two reasonable *constant* thresholds for initiating and relaxing social distancing. Part (a) of Figure S3 projects pandemic waves and lock-down periods based on an arbitrary threshold that is below the optimal constant threshold shown in Figure S1c and Part (b) similarly considers an arbitrary threshold above the optimal value.

Finally, we consider a scenario in which transmission is more effectively mitigated during relaxation of social distancing. When lock-downs are lifted, the population continues to reduce transmission by 80% rather than 40%. Figure S4 projects COVID-19 under a policy that has been optimized under this more optimistic scenario. The model projects that hospitalizations will remain under capacity without requiring another lock-down before September 2021 because of the stringent mitigation even during the relaxation period.

Table S1 is the analog of Table 1 from the main text, except that it reports values for projected days of lock-down, probabilities of exceeding hospital capacity, and COVID-19 mortality under the policies for Figure 1(a) and 1(b) of the main document. Tables S2 and S3 similarly report details for Figures S1a-S1b and S1c-S1d, respectively. Table S4 does so for Figures S3a-S3b, and Table S5 finally repeats these details for Figures S2 and S4.

**Fig. S1.**
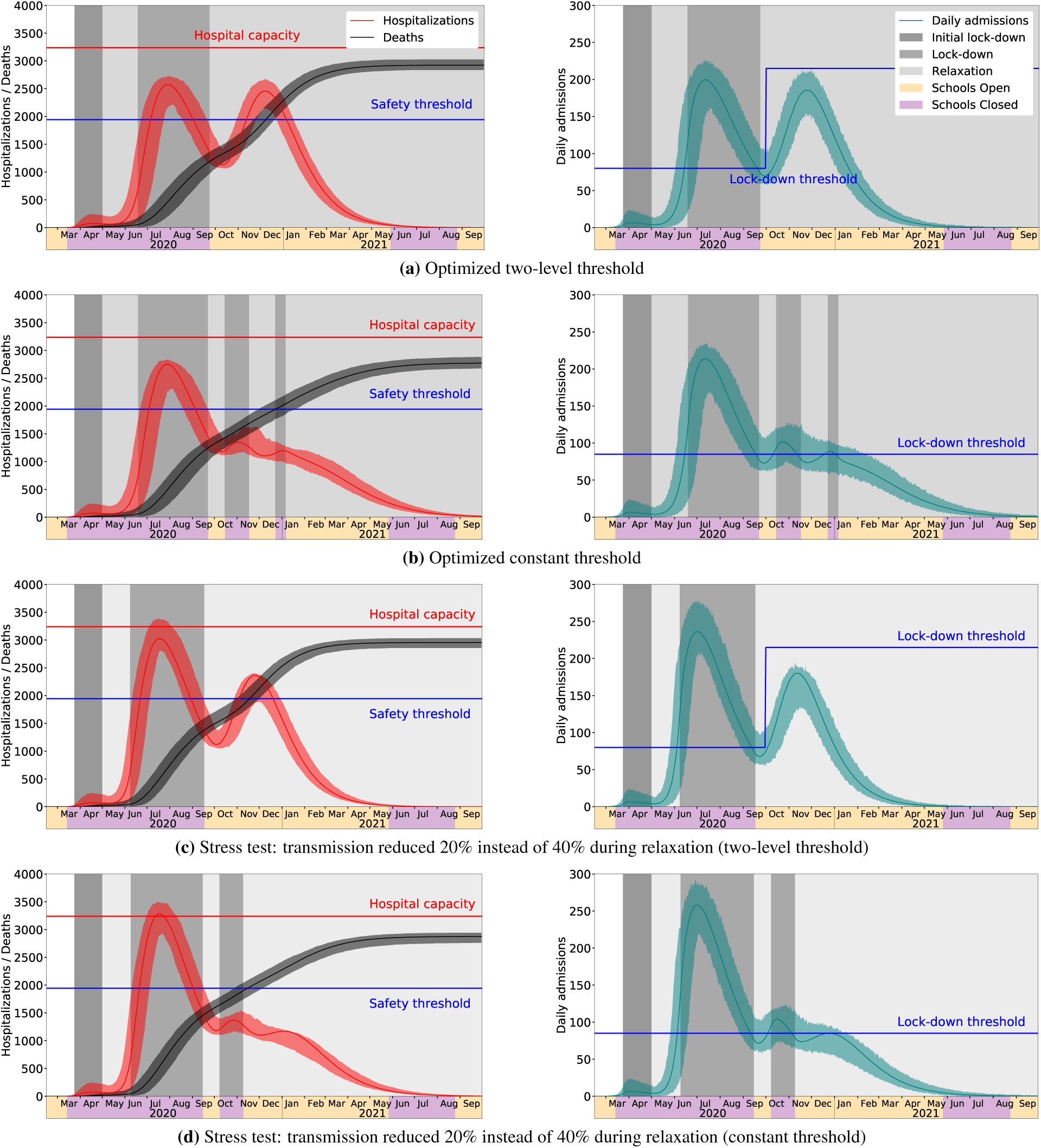
Stress test of optimized strategies with respect to transmission rates following the relaxation of social distancing. The graphs project COVID-19 hospitalizations and deaths in the Austin, Texas metropolitan area through September of 2021 under optimized policies for initiating and relaxing social distancing measures, assuming 95% effective cocooning of vulnerable populations. Left-hand panels show daily hospitalizations and cumulative deaths. COVID-19 surge capacity in Austin is approximately 3240 beds (red line). Right-hand panels show daily hospital admissions. Lock-downs are enacted when daily admissions (seven-day rolling average) surpass the indicated thresholds and are relaxed when admissions decline below the thresholds, if hospitalizations are below a safety threshold 60% capacity (blue line, left graphs). Part (a) repeats a figure presented in the main text showing the optimal two-step threshold. Part (b) instead uses a constant threshold. Parts (c) and (d) implement the policies optimized in parts (a) and (b), respectively. However, they assume that the public adheres only to 20% social distancing rather than the 40% for which the policy was designed. Hospital capacity may be exceeded if transmission rates are higher than expected during periods of relaxation. In all graphs, solid curves correspond to the point forecast and shaded regions give 90% prediction intervals based on 300 stochastic simulations.

**Fig. S2.**
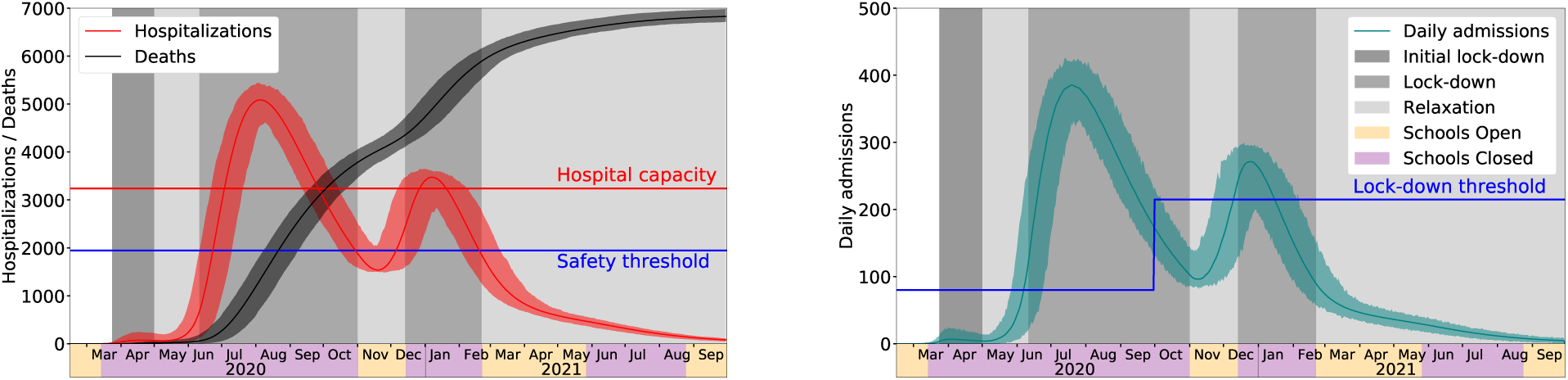
Stress test of optimized strategies with respect to the efficacy of cocooning vulnerable populations. The graphs project COVID-19 hospitalizations and deaths in the Austin, Texas metropolitan area through September of 2021 under optimized policies for initiating and relaxing social distancing measures. The left-hand panel show daily hospitalizations and cumulative deaths. COVID-19 surge capacity in Austin is approximately 3240 beds (red line). Lock-downs can only relax if the total hospitalizations are below a safety threshold of 60% capacity (blue line). The right-hand panel show daily hospital admissions. Lockdowns are enacted when daily admissions (seven-day rolling average) surpass the indicated thresholds and are relaxed when admissions decline below the thresholds, if hospitalizations are below 60% capacity. To make these graphs, we first derived the optimal thresholds for the baseline scenario which toggles between 90% (lock-down) and 40% (relaxation) transmission reduction, while assuming that cocooning remains 95% effective. Then, we then evaluate performance of the triggering policy under the assumption that cocooning is only 80% effective in reducing infection risk in vulnerable populations. Imperfect cocooning is projected to result in catastrophic health care surges and more than double the mortality. In both graphs, solid curves correspond to the point forecast and shaded regions give 90% prediction intervals based on 300 stochastic simulations.

**Fig. S3.**
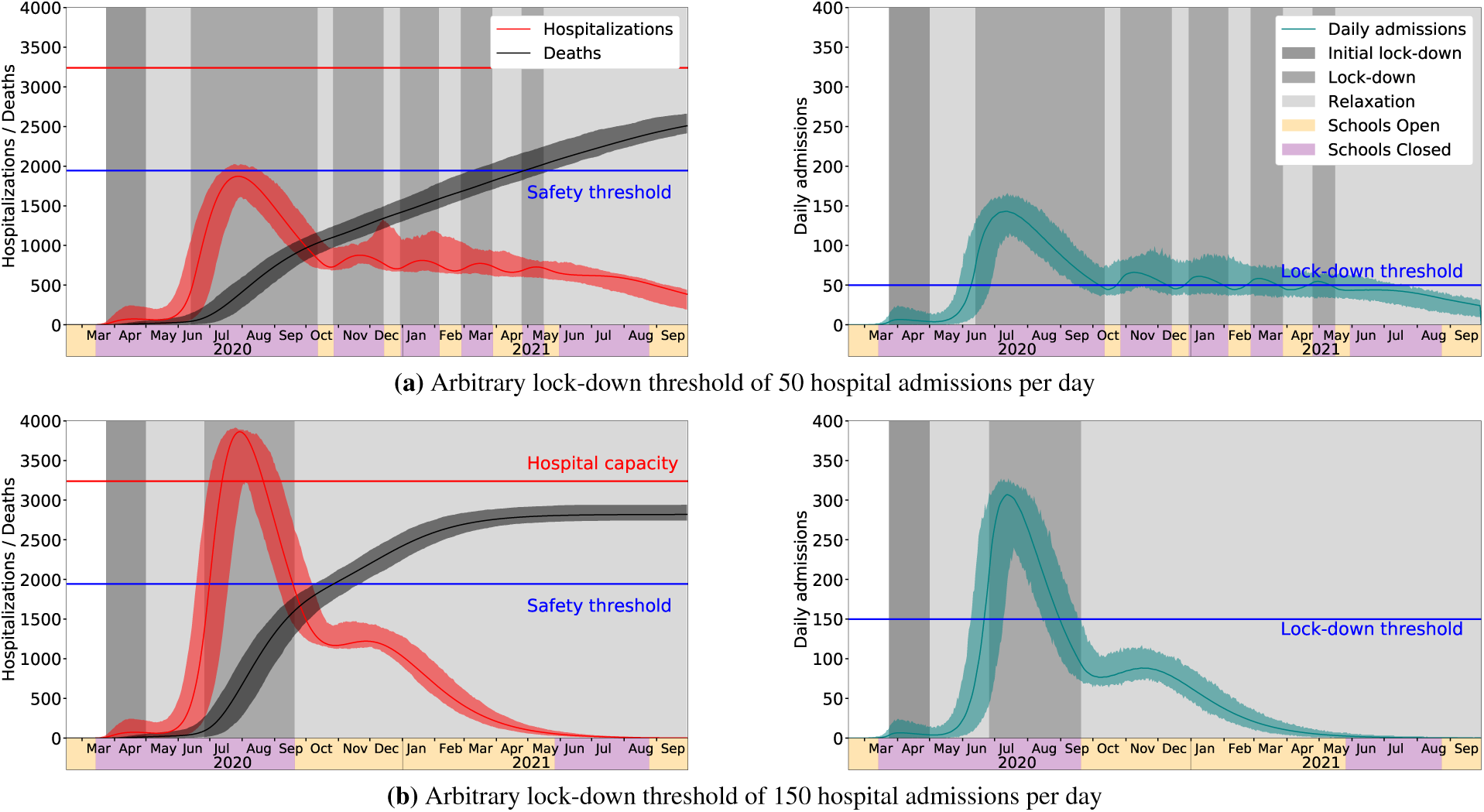
Non-optimized thresholds for triggering COVID-19 lock-downs. The graphs project COVID-19 hospitalizations and deaths in the Austin, Texas metropolitan area through September of 2021 under arbitrarily chosen triggers for initiating and relaxing social distancing measures, assuming 95% effective cocooning of vulnerable populations. Left-hand panels show daily hospitalizations and cumulative deaths. COVID-19 surge capacity in Austin is approximately 3240 beds (red line). Lock-downs can only relax if the total hospitalizations are below a safety threshold of 60% capacity (blue line). Right-hand panels show daily hospital admissions. Lock-downs are enacted when daily admissions (seven-day rolling average) surpass the indicated thresholds and are relaxed when admissions decline below the thresholds, if hospitalizations are below 60% capacity. Parts a and b assume arbitrary triggers that are below and above the optimal constant threshold from Figure S1c, respectively. In all graphs, solid curves correspond to the point forecast and shaded regions give 90% prediction intervals based on 300 stochastic simulations.

**Fig. S4.**
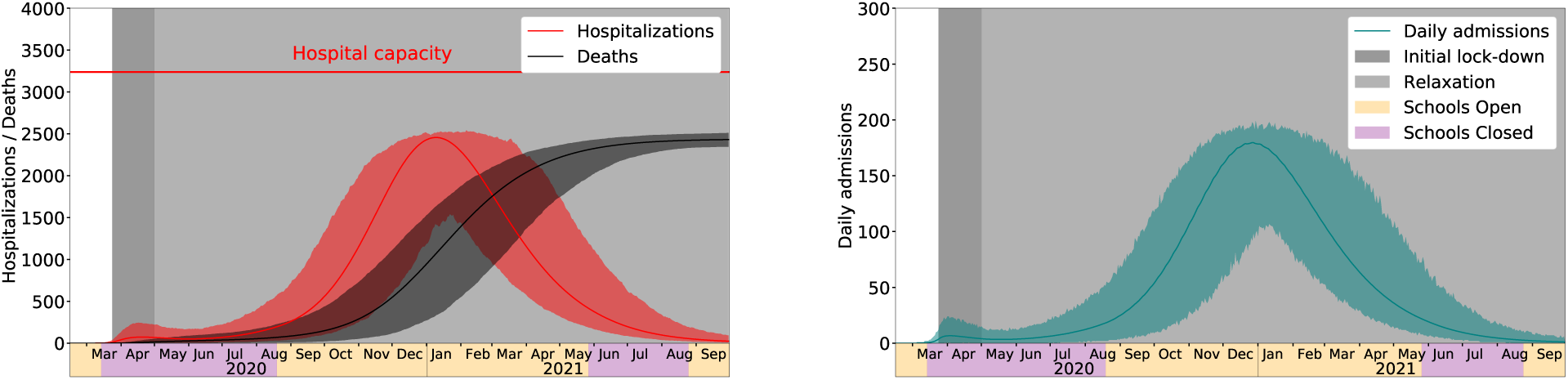
Projections assuming an 80% rather than 40% reduction in transmission during relaxation periods. The graphs project COVID-19 hospitalizations and deaths in the Austin, Texas metropolitan area through September of 2021 under optimized policies for initiating and relaxing social distancing measures. The left-hand panel show daily hospitalizations and cumulative deaths. If Austin maintains 80% reduction in transmission throughout the relaxation period, then hospitalizations will remain the local COVID-19 surge capacity of approximately 3240 beds (red line) without requiring a lock-down prior to September 2021. The right-hand panel show daily hospital admissions. In both graphs, solid curves correspond to the point forecast and shaded regions give 90% prediction intervals based on 300 stochastic simulations.

**Table S1:** Projected days of lock-down, probabilities of exceeding hospital capacity and COVID-19 mortality under the indefinite lock-down and indefinite relaxation. Companion of Figure 1(a) and 1(b) of the main document.

| Scenario |  | Indefinite lock-down | Indefinite relaxation |
| --- | --- | --- | --- |
| Days of lock-down | Mean | 555 | 38 |
|  | Median | 555 | 38 |
|  | 5%-95% PI | (555-555) | (38-38) |
| Cumulative deaths by Sep 30, 2021 | Mean | 81 | 2957 |
|  | Median | 65 | 2956 |
|  | 5%-95% PI | (10-202) | (2868-3040) |
| Risk Group | Age Group | % deaths |  |
| Low risk | 0-4y | 0.00 | 0.03 |
|  | 5-17y | 0.00 | 0.20 |
|  | 18-49y | 6.17 | 9.13 |
|  | 50-64y | 11.11 | 18.67 |
|  | 65y+ | 4.94 | 4.43 |
| High risk | 0-4y | 0.00 | 0.00 |
|  | 5-17y | 0.00 | 0.10 |
|  | 18-49y | 7.41 | 5.95 |
|  | 50-64y | 30.86 | 26.24 |
|  | 65y+ | 39.51 | 35.24 |
| Probability of exceeding % of hospital capacity | 60% | 0.000 | 1.000 |
|  | 70% | 0.000 | 1.000 |
|  | 80% | 0.000 | 1.000 |
|  | 90% | 0.000 | 1.000 |
|  | 100% | 0.000 | 1.000 |

**Table S2:**
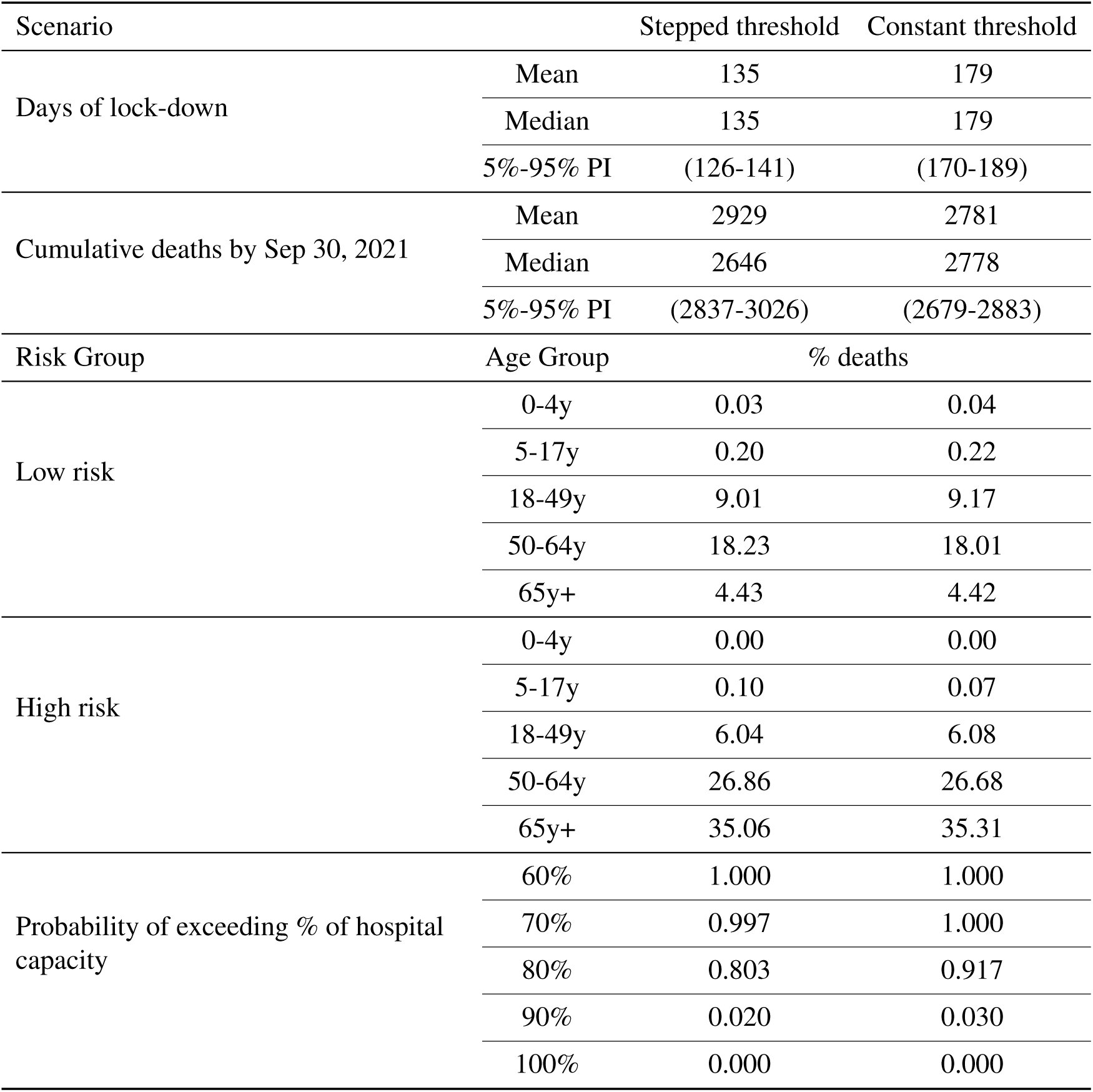
Projected days of lock-down, probabilities of exceeding hospital capacity and COVID-19 mortality under the optimized stepped threshold and the optimized constant threshold. These correspond to Figures S1a and S1b in which relaxed social distancing corresponds to a 40% reduction in transmission.

**Table S3:** Projected days of lock-down, probabilities of exceeding hospital capacity and COVID-19 mortality under stress tests for the stepped threshold and the constant threshold. These correspond to Figures S1c and S1d. Policies are trained assuming that relaxed social distancing corresponds to a 40% reduction in transmission, but then tested when relaxed social distancing instead corresponds to a 20% reduction in transmission.

| Scenario |  | Stepped threshold | Constant threshold |
| --- | --- | --- | --- |
| Days of lock-down | Mean | 139 | 173 |
|  | Median | 139 | 173 |
|  | 5%-95% PI | (136-144) | (165-185) |
| Cumulative deaths by Sep 30, 2021 | Mean | 2950 | 2852 |
|  | Median | 2948 | 2855 |
|  | 5%-95% PI | (2857-3033) | (2761-2942) |
| Risk Group | Age Group | % deaths |  |
| Low risk | 0-4y | 0.03 | 0.04 |
|  | 5-17y | 0.20 | 0.21 |
|  | 18-49y | 9.05 | 9.15 |
|  | 50-64y | 18.27 | 18.16 |
|  | 65y+ | 4.44 | 4.45 |
| High risk | 0-4y | 0.00 | 0.00 |
|  | 5-17y | 0.10 | 0.11 |
|  | 18-49y | 6.00 | 5.96 |
|  | 50-64y | 26.68 | 26.61 |
|  | 65y+ | 35.22 | 35.31 |
| Probability of exceeding % of hospital capacity | 70% | 1.00 | 1.000 |
|  | 80% | 0.997 | 0.997 |
|  | 90% | 0.983 | 0.997 |
|  | 100% | 0.437 | 0.797 |
|  | 110% | 0.003 | 0.053 |

**Table S4:** Projected days of lock-down, probabilities of exceeding hospital capacity and COVID-19 mortality under non-optimized constant thresholds. These correspond to Figures S3a and S3b. Relaxed social distancing corresponds to a 40% reduction in transmission.

| Scenario |  | Threshold of 50 | Threshold of 150 |
| --- | --- | --- | --- |
| Days of lock-down | Mean | 291 | 123 |
|  | Median | 292 | 124 |
|  | 5%-95% PI | (270-308) | (121-125) |
| Cumulative deaths by Sep 30, 2021 | Mean | 2526 | 2839 |
|  | Median | 2520 | 2837 |
|  | 5%-95% PI | (2416-2660) | (2744-2940) |
| Risk Group | Age Group | % deaths |  |
| Low risk | 0-4y | 0.04 | 0.04 |
|  | 5-17y | 0.20 | 0.21 |
|  | 18-49y | 9.34 | 9.16 |
|  | 50-64y | 17.93 | 18.07 |
|  | 65y+ | 4.43 | 4.44 |
| High risk | 0-4y | 0.00 | 0.00 |
|  | 5-17y | 0.08 | 0.11 |
|  | 18-49y | 6.02 | 6.02 |
|  | 50-64y | 26.72 | 26.70 |
|  | 65y+ | 35.23 | 35.26 |
| Probability of exceeding % of hospital capacity | 60% | 0.293 | 1.000 |
|  | 70% | 0.000 | 1.000 |
|  | 80% | 0.000 | 1.000 |
|  | 90% | 0.000 | 1.000 |
|  | 100% | 0.000 | 1.000 |

**Table S5:** Projected days of lock-down, probabilities of exceeding hospital capacity and COVID-19 mortality under two scenarios. The first is a stress test, which optimizes assuming cocooning is 95% effective, but tests when it is instead 80% effective; this corresponds to Figure S2. The second policy corresponds to relaxed social distancing corresponding to 80% (rather than our nominal 40%) reduction in transmission under cocooning, which is 95% effective. In this case we do not need to enforce a lock-down after May 1, 2020 to prevent hospitalizations from exceeding capacity. The right-most column corresponds to Figure S4.

| Scenario |  | 80% Cocooning | 80% Relaxed |
| --- | --- | --- | --- |
| Days of lock-down | Mean | 249 | 38 |
|  | Median | 249 | 38 |
|  | 5%-95% PI | (243-254) | (38-38) |
| Cumulative deaths by Sep 30, 2021 | Mean | 6839 | 2426 |
|  | Median | 6830 | 2427 |
|  | 5%-95% PI | (6715-6977) | (2346-2511) |
| Risk Group | Age Group | % deaths |  |
| Low risk | 0-4y | 0.04 | 0.04 |
|  | 5-17y | 0.20 | 0.21 |
|  | 18-49y | 9.34 | 8.90 |
|  | 50-64y | 17.93 | 16.61 |
|  | 65y+ | 4.43 | 4.45 |
| High risk | 0-4y | 0.00 | 0.00 |
|  | 5-17y | 0.08 | 0.12 |
|  | 18-49y | 6.02 | 6.43 |
|  | 50-64y | 26.72 | 28.03 |
|  | 65y+ | 35.23 | 35.20 |
| Probability of exceeding % of hospital capacity | 60% | 1.000 | 0.997 |
|  | 70% | 1.000 | 0.993 |
|  | 80% | 1.000 | 0.080 |
|  | 90% | 1.000 | 0.000 |
|  | 100% | 1.000 | 0.000 |

### Model Parameters

#### Age-stratified proportion of population at high-risk for COVID-19 complications

We estimate age-specific proportions of the population at high risk of complications from COVID-19 based on data for Austin, TX and Round-Rock, TX from the CDC’s 500 cities project (Figure S5) (*21*). We assume that high risk conditions for COVID-19 are the same as those specified for influenza by the CDC (*22*). The CDC’s 500 cities project provides city-specific estimates of prevalence for several of these conditions among adults (*23*). The estimates were obtained from the 2015–2016 Behavioral Risk Factor Surveillance System (BRFSS) data using a small-area estimation methodology called multi-level regression and poststratification (*24,25*). It links geocoded health surveys to high spatial resolution population demographic and socioeconomic data (*25*).

**Fig. S5.**
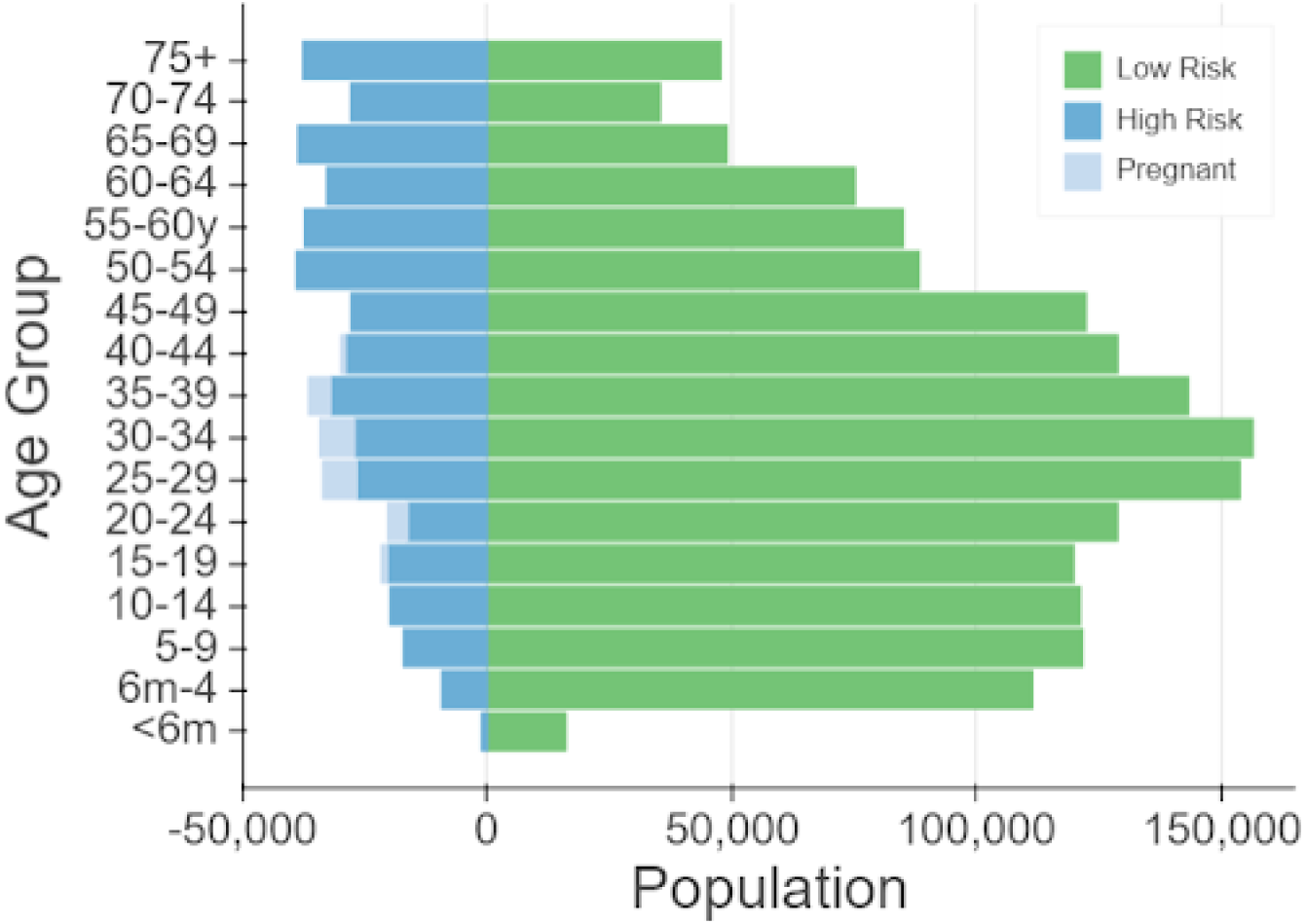
Demographic and risk composition of the Austin-Round Rock MSA. Bars indicate age-specific population sizes, separated by low risk, high risk, and pregnant women. High risk is defined as individuals with cancer, chronic kidney disease, chronic obstructive pulmonary disease (COPD), heart disease, stroke, asthma, diabetes, HIV/AIDS, and morbid obesity.

#### High-risk proportions for adults

To estimate the proportion of adults at high risk for complications, we use the CDC’s 500 cities data mentioned above, as well as data on the prevalence of HIV/AIDS, obesity and pregnancy among adults (Table S13).

The CDC 500 cities dataset includes the prevalence of each condition on its own, rather than the prevalence of multiple conditions (e.g., dyads or triads). Thus, we use separate co-morbidity estimates to determine overlap. The following reference about chronic conditions (*26*) gives US estimates for the proportion of the adult population with 0, 1 or 2+ chronic conditions, per age group. Using this and the 500 cities data we can estimate the proportion of the population *p_HR_* in each age group in each city with at least one chronic condition listed in the CDC 500 cities data (Table S13) putting them at high-risk for flu complications.

#### HIV

We use the data from Table 20a in CDC HIV surveillance report (*27*) to estimate the population in each risk group living with HIV in the US (last column, 2015 data). Assuming independence between HIV and other chronic conditions, we increase the proportion of the population at high-risk for influenza to account for individuals with HIV but no other underlying conditions.

#### Morbid obesity

A BMI over 40kg/m^2^ indicates morbid obesity, and is considered high risk for influenza. The 500 Cities Project reports the prevalence of obese people in each city with BMI over 30kg/m2 (not necessarily morbid obesity). We use the data from Table 1 in (*28*) to estimate the proportion of people with BMI > 30 that actually have BMI> 40 (across the US); we then apply this to the 500 Cities obesity data to estimate the proportion of people who are morbidly obese in each city. Table 1 of (*28*) suggests that 51.2% of morbidly obese adults have at least one other high risk chronic condition, and update our high-risk population estimates accordingly to account for overlap.

#### Pregnancy

We separately estimate the number of pregnant women in each age group and each city, following the methodology in a CDC reproductive health report (*29*). We assume independence between any of the high-risk factors and pregnancy, and further assume that half the population are women; also, see (*30*).

#### Estimating high-risk proportions for children

Since the 500 Cities Project only reports data for adults 18 years and older, we take a different approach to estimating the proportion of children at high risk for severe influenza. The two most prevalent risk factors for children are asthma and obesity; we also account for childhood diabetes, HIV and cancer. From (*31*), we obtain national estimates of chronic conditions in children. For asthma, we assume that variation among cities will be similar for children and adults. Thus, we use the relative prevalence of asthma in adults to scale our estimates for children in each city. The prevalence of HIV and cancer in children are taken from CDC HIV surveillance report (*27*) and cancer research report (*32*), respectively.

We first estimate the proportion of children having either asthma, diabetes, cancer or HIV (assuming no overlap in these conditions). We estimate city-level morbid obesity in children using the estimated morbid obesity in adults multiplied by a national constant ratio for each age group estimated from (*33*), this ratio represents the prevalence in morbid obesity in children given the one observed in adults. From (*34*), we estimate that 25% of morbidly obese children have another high-risk condition and adjust our final estimates accordingly.

#### Resulting estimates

We compare our estimates for the Austin-Round Rock Metropolitan Area to published national-level estimates (*35*) of the proportion of each age group with underlying high risk conditions (Table S13). The biggest difference is observed in older adults, with Austin having a lower proportion at risk for complications for COVID-19 than the national average; for 25–39 year olds the high risk proportion is slightly higher than the national average.

**Table S6:** Initial conditions, school closures and social distancing policies

| Variable | Settings |
| --- | --- |
| Initial day of simulation | 2/15/2020 |
| Initial infection number in locations | 1 symptomatic case in 18-49 year age group |
| Social distancing reduction in contacts | As marked in the corresponding table and figure. Note that social distancing is determined from the triggers. |
| Age-specific and day-specific contact rates | Home, work, other and school matrices provided in Tables S9-S12<br>As marked in corresponding table and figures with social level distance denoted as SD and cocooning level denoted as CO:<br>If no measure is in place (other than social distance):<br>Weekday = $(1-SD)$ (home + school + work + other)<br>Weekend = $(1-SD)$ (home + other)<br>If only schools are closed:<br>Weekday = $(1-SD)$ (home + work + other)<br>Weekend = $(1-SD)$ (home + other)<br>If only cocooning is on:<br>Low risk and age groups 1-4<br>Weekday = $(1-SD)$ (home + school + work + other)<br>Weekend = $(1-SD)$ (home + other)<br>High risk and age group 5<br>Weekday = $(1-CO)$ (home + school + work + other)<br>Weekend = $(1-CO)$ (home + other)<br>If schools are closed and cocooning is on:<br>Low risk and age groups 1-4<br>Weekday = $(1-SD)$ (home + work + other)<br>Weekend = $(1-SD)$ (home + other)<br>High risk and age group 5<br>Weekday = $(1-CO)$ (home + work + other)<br>Weekend = $(1-CO)$ (home + other) |

**Table S7:**
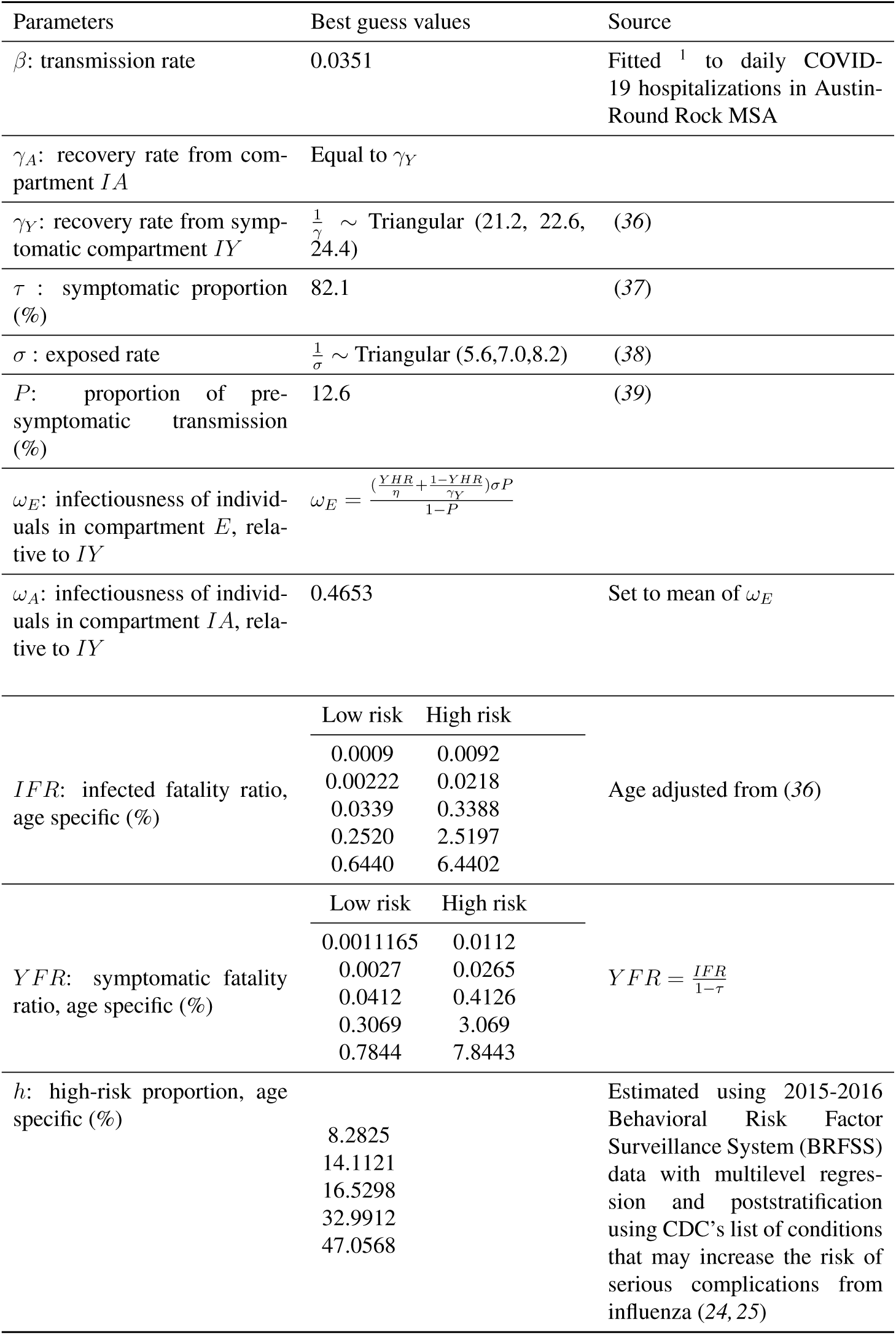
Model parameters

**Table S8:**
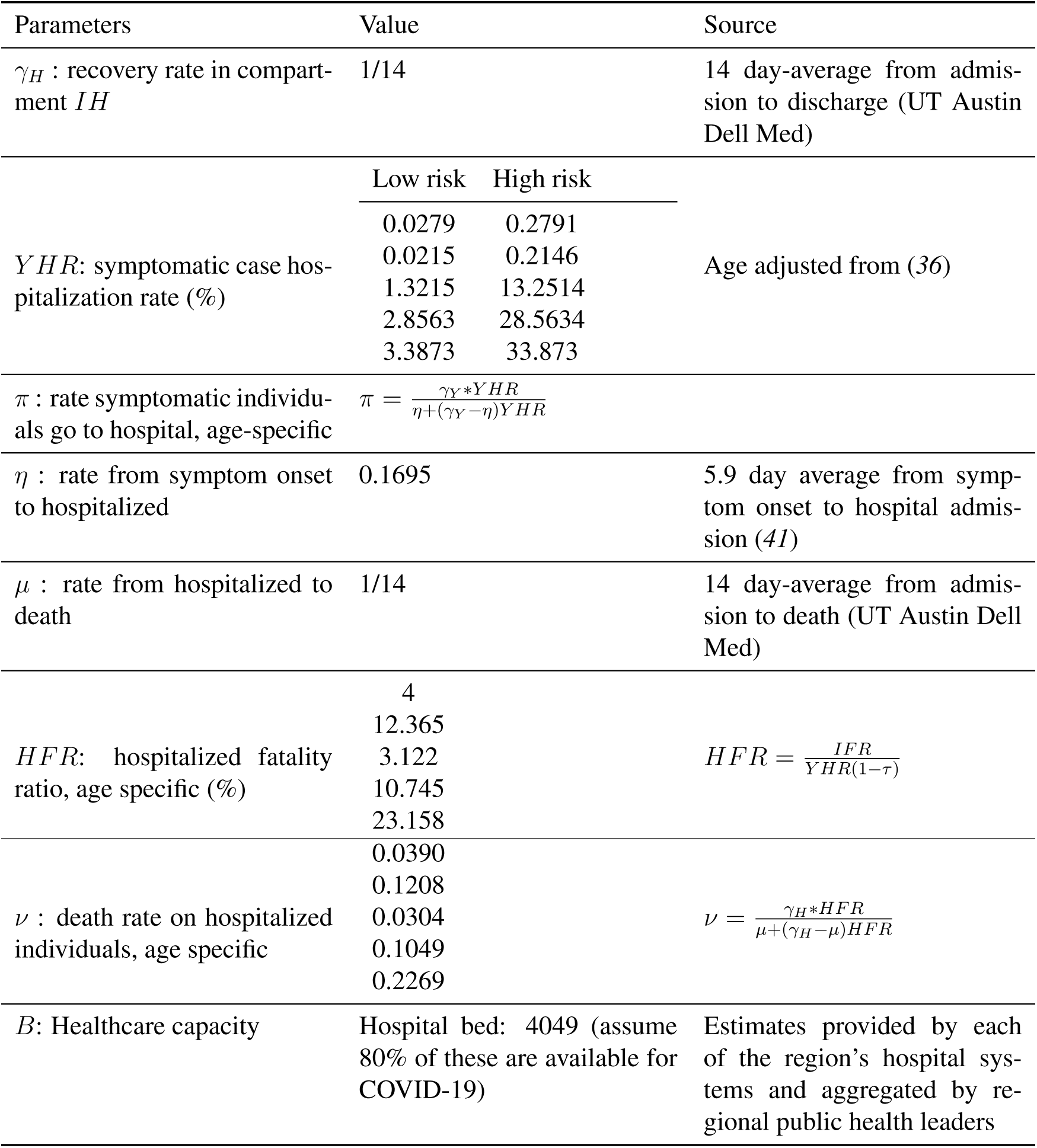
Hospitalization parameters

**Table S9:** Home contact matrix. Daily number contacts by age group at home.

|  | 0-4y | 5-17y | 18-49y | 50-64y | 65y+ |
| --- | --- | --- | --- | --- | --- |
| 0-4y | 0.5 | 0.9 | 2.0 | 0.1 | 0.0 |
| 5-17y | 0.2 | 1.7 | 1.9 | 0.2 | 0.0 |
| 18-49y | 0.2 | 0.9 | 1.7 | 0.2 | 0.0 |
| 50-64y | 0.2 | 0.7 | 1.2 | 1.0 | 0.1 |
| 65y+ | 0.1 | 0.7 | 1.0 | 0.3 | 0.6 |

**Table S10:** School contact matrix. Daily number contacts by age group at school.

|  | 0-4y | 5-17y | 18-49y | 50-64y | 65y+ |
| --- | --- | --- | --- | --- | --- |
| 0-4y | 1.0 | 0.5 | 0.4 | 0.1 | 0.0 |
| 5-17y | 0.2 | 3.7 | 0.9 | 0.1 | 0.0 |
| 18-49y | 0.0 | 0.7 | 0.8 | 0.0 | 0.0 |
| 50-64y | 0.1 | 0.8 | 0.5 | 0.1 | 0.0 |
| 65y+ | 0.0 | 0.0 | 0.1 | 0.0 | 0.0 |

**Table S11:** Work contact matrix. Daily number contacts by age group at work.

|  | 0-4y | 5-17y | 18-49y | 50-64y | 65y+ |
| --- | --- | --- | --- | --- | --- |
| 0-4y | 0.0 | 0.0 | 0.0 | 0.0 | 0.0 |
| 5-17y | 0.0 | 0.1 | 0.4 | 0.0 | 0.0 |
| 18-49y | 0.0 | 0.2 | 4.5 | 0.8 | 0.0 |
| 50-64y | 0.0 | 0.1 | 2.8 | 0.9 | 0.0 |
| 65y+ | 0.0 | 0.0 | 0.1 | 0.0 | 0.0 |

**Table S12:** Others contact matrix. Daily number contacts by age group at other locations.

|  | 0-4y | 5-17y | 18-49y | 50-64y | 65y+ |
| --- | --- | --- | --- | --- | --- |
| 0-4y | 0.7 | 0.7 | 1.8 | 0.6 | 0.3 |
| 5-17y | 0.2 | 2.6 | 2.1 | 0.4 | 0.2 |
| 18-49y | 0.1 | 0.7 | 3.3 | 0.6 | 0.2 |
| 50-64y | 0.1 | 0.3 | 2.2 | 1.1 | 0.4 |
| 65y+ | 0.0 | 0.2 | 1.3 | 0.8 | 0.6 |

**Table S13:** High-risk conditions for influenza and data sources for prevalence estimation

| Condition | Data source |
| --- | --- |
| Cancer (except skin) | CDC 500 cities (22) |
| Chronic kidney disease | CDC 500 cities (22) |
| COPD | CDC 500 cities (22) |
| Coronary heart disease | CDC 500 cities (22) |
| Stroke | CDC 500 cities (22) |
| Asthma | CDC 500 cities (22) |
| Diabetes | CDC 500 cities (22) |
| HIV/AIDS | CDC HIV Surveillance Report (27) |
| Obesity | CDC 500 cities and (28, 34) |
| Pregnancy | National Vital Statistics Reports (42) and abortion data (43) |

**Table S14:** Comparison between published national estimates and Austin-Round Rock MSA estimates of the percent of the population at high-risk of influenza/COVID-19 complications.

| Age Group | National estimates (33) | Austin<br>(excluding pregnancy) | Pregnant women<br>(proportion of age group) |
| --- | --- | --- | --- |
| 0-0.5y | n/a | 6.8 | - |
| 0.5-4y | 6.8 | 7.4 | - |
| 5-9 y | 11.7 | 11.6 | - |
| 10-14y | 11.7 | 13.0 | - |
| 15-19y | 11.8 | 13.3 | 1.7 |
| 20-24y | 12.4 | 10.3 | 5.1 |
| 25-34y | 15.7 | 13.5 | 7.8 |
| 35-39y | 15.7 | 17.0 | 5.1 |
| 40-44y | 15.7 | 17.4 | 1.2 |
| 45-49y | 15.7 | 17.7 | - |
| 50-54y | 30.6 | 29.6 | - |
| 55-60y | 30.6 | 29.5 | - |
| 60-64y | 30.6 | 29.3 | - |
| 65-69y | 47.0 | 42.2 | - |
| 70-74y | 47.0 | 42.2 | - |
| 75y+ | 47.0 | 42.2 | - |

